# Impact of Trunk Function and Lower Limb Paralysis on Independence in Activities of Daily Living in Patients with Stroke

**DOI:** 10.1101/2024.12.02.24318305

**Authors:** Takato Nishida, Shun Sawai, Shoya Fujikawa, Ryosuke Yamamoto, Yusuke Shizuka, Takayuki Maru, Kotaro Nakagawa, Hideki Nakano

**Affiliations:** Department of Physical Therapy, Faculty of Rehabilitation and Care, Seijoh University, Tokai, Japan; Graduate School of Health Sciences, Kyoto Tachibana University, Kyoto, Japan; Department of Rehabilitation, Kyoto Kuno Hospital, Kyoto, Japan; Department of Rehabilitation, Tesseikai Neurosurgical Hospital, Shijonawate, Japan; Department of Physical Therapy, Faculty of Health Sciences, Kyoto Tachibana University, Kyoto, Japan; Department of Rehabilitation, Junshinkai Kobe Hospital, Kobe, Japan; Nagashima Neurosurgery Rehabilitation Clinic, Osaka, Japan

**Keywords:** trunk function, lower limb paralysis, ADL, stroke, hierarchical regression analysis

## Abstract

Stroke is a major global health issue, and many patients experience motor paralysis and sensory impairments that affect their independence in activities of daily living (ADL). Trunk and lower limb functions are crucial in post-stroke ADL independence. Although these two functions are closely related, few studies have evaluated them in combination, and the importance of assessment methods that consider their mutual relationship has not been thoroughly examined. In this study, we aimed to clarify the degree to which trunk function and lower limb paralysis impact ADL independence when evaluated individually versus in combination, through a hierarchical regression analysis, and to verify the significance of the combined assessment of both functions. This cross-sectional study included 51 patients with first-ever stroke and hemiplegia. Trunk function was assessed using the Trunk Impairment Scale, lower limb paralysis was evaluated using the Brunnstrom Recovery Stage for the lower extremities, and ADL independence was measured using the Functional Independence Measure. Hierarchical regression analysis was conducted to examine the impact of trunk and lower limb functions on ADL independence. Across two regression models, the assessment of trunk and lower limb function in combination significantly improved the accuracy in reflecting ADL independence compared with the assessment of each function individually. The findings suggest that a combined assessment of both trunk and lower limb functions is a valuable evaluation method in the rehabilitation of patients with stroke.

## 1 Introduction

Stroke, the second most common cause of death and third most common cause of disability, is a leading global health issue(Du et al., 2023; Zhang et al., 2024). Between 1990 and 2019, the incidence and prevalence of stroke increased by 18.5% and 32%, respectively. In 2019 alone, approximately 6.1 million people died from stroke worldwide, highlighting its significant health burden (Zhang et al., 2024). The particularly high incidence and mortality rates of stroke in low-income regions contribute to health disparities, emphasizing the need for global initiatives in stroke prevention and treatment (Feigin et al., 2022).

Stroke causes a variety of functional impairments, such as motor paralysis, sensory deficits, and speech disorders, which hinder the recovery of independence in activities of daily living (ADL) and walking ability (Patel et al., 2000). Improving ADL independence is a primary goal in post-stroke rehabilitation, and it requires the multidimensional assessment of functions. In particular, accurate evaluation of trunk function and lower limb paralysis is crucial for developing effective treatment programs (van der Putten et al., 1999; Hsieh et al., 2002; Meijer et al., 2003; Van Criekinge et al., 2017).

Trunk function plays multiple roles, including assisting with support against gravity, respiratory function, and postural control in response to both internal and external disturbances (Thijs et al., 2023). Impairments in trunk support and postural control significantly affect the recovery of ADL and walking ability in patients with stroke (Karatas et al., 2004; Van Criekinge et al., 2019).

Furthermore, acute-phase trunk function is strongly associated with independence in ADL 6 months after stroke onset, and the evaluation of trunk function can predict post-discharge balance and mobility abilities (Wang et al., 2005; Verheyden et al., 2006).

Similarly, lower limb function plays a critical role in ADL independence. Lower limb paralysis, weight-bearing on the non-paralyzed side, and lower limb muscle strength have been associated with walking ability and ADL independence in patients with stroke (Pradon et al., 2013; Matsuyama, 2018). This is particularly evident in ADL tasks that require anti-gravity activities, such as walking and maintaining a standing posture. Additionally, improvements in upper and lower limb paralysis have been linked to enhanced ADL independence (Kwakkel et al., 2002; Liu and Liu, 2022). A decline in lower limb function, including motor paralysis, poses a serious challenge to walking and ADL independence in patients with stroke. As such, the evaluation of lower limb function to facilitate ADL recovery is warranted. Both trunk and lower limb functions have been individually associated with ADL independence, and in ADL tasks that require significant standing activity, coordinated movements between the trunk and lower limbs are crucial (Sullivan et al., 2002).

Patients with stroke, who often experience motor impairments on one side of the body, may rely more heavily on the interrelationship between trunk and lower limb functions than do healthy individuals.

However, most previous studies have evaluated trunk and lower limb functions separately, and comprehensive assessment methods that consider their interrelationship have been insufficiently investigated (Sullivan et al., 2002). Additionally, some studies focusing on the impact of trunk function on ADL independence did not include ADL tasks involving mobility, thereby limiting the practicality of the evaluations (Hsieh et al., 2002). To verify the importance of comprehensive assessment methods that account for the interaction between trunk and lower limb functions in stroke rehabilitation, the incremental addition of functional evaluation items is necessary, as well as the use of practical ADL measures to assess the impact on ADL independence.

Therefore, in this study, we aimed to clarify the extent to which trunk function and lower limb paralysis affect ADL independence, both when evaluated individually and in combination, through stepwise analysis. We also sought to verify the importance of a combined assessment method that integrates both functions.

## 2 Methods

### 2.1 Participants

This cross-sectional study included patients with first-time stroke and hemiplegia who were admitted to Aichi-Pref Saiseikai Rehabilitation Hospital between July 2017 and October 2018.

The exclusion criteria were subarachnoid hemorrhage, infratentorial lesions, the inability to comprehend verbal instructions, and lack of consent to participate in the study.

### 2.2 Ethics Declarations

This study was conducted in accordance with the Declaration of Helsinki and informed consent was obtained from all participants. This study was approved by the Research Ethics Committee of Seijoh University (Approval number: 2016C0035; Approval date: July 12, 2017) and the Aichi-Pref Saiseikai Rehabilitation Hospital (Approval number: 201705; Approval date: March 24, 2017). This study was registered in the UMIN Clinical Trials Registry (UMIN000056341).

### 2.3 Measures

We collected data on age, sex, stroke type, paralyzed side, and days since stroke onset from medical records as basic information, and administered the Brunnstrom Recovery Stage for the lower extremities (BRS-LE) to assess motor paralysis, Trunk Impairment Scale (TIS) to assess trunk function, and Functional Independence Measure (FIM) to assess ADL independence. The BRS-LE and TIS were administered on the same day by the principal investigator. FIM scores were collected from assessments conducted by the responsible physical therapists, occupational therapists, and nurses during the same period.

#### 2.3.1 Brunnstrom Recover Stage for the lower extremities

The BRS-LE, proposed by Brunnstrom, evaluates motor paralysis as a qualitative phenomenon based on changes in movement patterns (Brunnstrom, 1966). It comprises assessments of the upper limbs, fingers, and lower limbs, with the degree of separation of associated and synergistic movements rated from Stage I to VI, where VI indicates the mildest form of motor paralysis. This tool was selected owing to its frequent use in Japan as well as its utility as a common language in research, as it serves as the basis for developing scales such as the Fugl–Meyer Assessment and Chedoke–McMaster Stroke Assessment (Fugl-Meyer et al., 1975; Gowland et al., 1993).

#### 2.3.2 Trunk Impairment Scale

The TIS, developed by Verheyden et al., is a clinical assessment tool designed to evaluate motor dysfunction in the trunk (Verheyden et al., 2004). It comprises 17 items (scored out of 23 points) covering static sitting balance, dynamic sitting balance, and coordination. Higher scores indicate better trunk motor function. We adopted the TIS as the trunk function evaluation in this study owing to its reported reliability and validity (Verheyden et al., 2004).

#### 2.3.3 Functional Independence Measure

The FIM measures practical ADL independence (Granger et al., 1993). It comprises 13 motor items and 5 cognitive items, with each item rated on a 7-point scale ranging from 1 (total assistance) to 7 (independence). The FIM is widely used in Japan’s recovery rehabilitation wards, where it is also employed in calculating medical fees. Herein, we used the total score for the motor items in the analysis.

### 2.4 Statistical Analysis

The Shapiro–Wilk test was used to confirm the normality of the data. We used hierarchical regression analysis to analyze the impact of motor paralysis and trunk function on ADL independence and the forced-entry method to examine variables that explained ADL independence. The dependent variable was the FIM score, and the independent variables were age, days since stroke onset, BRS-LE stage, and TIS score. In Step 1, age, days since stroke onset, and TIS score were entered, followed by BRS-LE stage in Step 2. To consider the possible influence of variable order, an alternate analysis was performed by entering age, days since stroke onset, and BRS-LE stage in Step 1, followed by TIS score in Step 2. All statistical analyses were conducted using SPSS Statistics, Version 29 (IBM Corp., Armonk, NY, United States), with the significance level set at 5%.

## 3 Results

### 3.1 Participant Characteristics

The analysis included the data of 51 participants. Table 1 shows the characteristics of the participants.

**Table 1.** Participant characteristics.

| Characteristics | N = 51 |
| --- | --- |
| Age (years) | 71.5 ± 13.3 |
| Sex (male/female) | 29/22 |
| Stroke type (ischemic/hemorrhagic) | 29/22 |
| Hemiplegic side (left/right) | 27/24 |
| Days since stroke onset (days) | 40 (28–57) |
| BRS-LE stage (I/II/III/IV/V/VI) | 4/8/2/9/6/22 |
| TIS score | 17 (7–20) |
| FIM motor score | 54 (21–77) |

Data are shown as mean ± standard deviation or median (interquartile range), as appropriate. BRS-LE: Brunnstrom Recovery Stage for the lower extremities; TIS: Trunk Impairment Scale; FIM: Functional Independence Measure

### 3.2 Hierarchical Multiple Regression Analysis using the TIS score

Table 2 shows the results of the hierarchical regression analysis using the TIS score as an independent variable and the total motor score of the FIM as the dependent variable. The variance inflation factor for all the variables was < 10.0, indicating no multicollinearity. In Step 1, age, days since stroke onset, and TIS score were entered into the model. The adjusted R^2^ of this model was 0.71, indicating that these variables had a certain explanatory power for the total motor score of the FIM. The unstandardized regression coefficient for each variable was as follows: age, β = -0.45 (p < 0.01); days since stroke onset, β = -0.05 (p = 0.51); and TIS score, β = 2.80 (p < 0.01). Age and the TIS score were the variables that significantly influenced the regression model. Subsequently, in Step 2, the model was tested by adding BRS-LE stage. The adjusted R^2^ of the Step 2 model was 0.75, similar to that of the Step 1 model, indicating a certain explanatory power for the total motor score of the FIM. Moreover, adding the BRS-LE stage significantly increased the change in F-value from the Step 1 model, indicating that the BRS-LE stage contributed to improving the model (ΔF = 8.81, p < 0.01).

**Table 2.** Hierarchical multiple regression analysis using TIS score.

| Step and variables | $\beta$ | SE | 95% CI | p | Adjusted R<sup>2</sup> | $\Delta F$ |
| --- | --- | --- | --- | --- | --- | --- |
| Step 1 | - | - | - | - | 0.71 | 42.45 |
| Age | -0.45 | 0.16 | -0.77, -0.12 | <0.01 | - | - |
| Days since stroke onset | -0.05 | 0.07 | -0.19, 0.10 | 0.51 | - | - |
| TIS score | 2.80 | 0.27 | 2.26, 3.34 | <0.01 | - | - |
| Step 2 | - | - | - | - | 0.75 | 8.81* |
| Age | -0.39 | 0.15 | -0.69, -0.08 | 0.01 | - | - |
| Days since stroke onset | -0.02 | 0.07 | -0.16, 0.11 | 0.73 | - | - |
| TIS score | 1.24 | 0.58 | 0.07, 2.41 | 0.04 | - | - |
| BRS-LE stage | 8.11 | 2.73 | 2.61, 13.61 | <0.01 | - | - |

\*p < 0.01. $\beta$ : unstandardized coefficient; SE: standard error; CI: confidence interval; Adjusted R<sup>2</sup>: adjusted coefficient of determination; $\Delta F$ : change in F-value from one model to another

### 3.3 Hierarchical Multiple Regression Analysis using the BRS-LE stage

Table 3 shows the results of the hierarchical regression analysis using the BRS-LE stage as an independent variable and the total motor score of the FIM as the dependent variable. The variance inflation factor for all the variables was < 10.0, indicating no multicollinearity. In Step 1, age, days since stroke onset, and BRS-LE stage were entered into the model. The adjusted R^2^ of this model was 0.74, indicating that these variables had a certain explanatory power for the total motor score of the FIM. The unstandardized regression coefficient for each variable was as follows: age, β = -0.36 (p = 0.03); days since stroke onset, β = -0.02 (p = 0.77); and BRS-LE stage, β = 13.37 (p < 0.01). Age and the BRS-LE stage were the variables that significantly influenced the regression model.

**Table 3.** Hierarchical multiple regression analysis with inputs from BRS-LE.

| Step and variables | $\beta$ | SE | 95%CI | p | Adjusted R<sup>2</sup> | $\Delta F$ |
| --- | --- | --- | --- | --- | --- | --- |
| Step 1 | - | - | - | - | 0.74 | 47.36 |
| Age | -0.36 | 0.16 | -0.67, -0.04 | 0.03 | - | - |
| Days since stroke onset | -0.02 | 0.07 | -0.16, 0.12 | 0.77 | - | - |
| BRS-LE stage | 13.37 | 1.21 | 10.93, 15.81 | <0.01 | - | - |
| Step 2 | - | - | - | - | 0.75 | 4.55* |
| Age | -0.39 | 0.15 | -0.69, -0.08 | 0.01 | - | - |
| Days since stroke onset | -0.02 | 0.07 | -0.16, 0.11 | 0.73 | - | - |
| BRS-LE stage | 8.11 | 0.50 | 2.61, 13.61 | <0.01 | - | - |
| TIS score | 1.24 | 0.36 | 0.07, 2.41 | 0.04 | - | - |

\*p < 0.01

Subsequently, in Step 2, the model was tested by adding the TIS score. The adjusted R^2^ of the Step 2 model was 0.75, similar to that of the Step 1 model, indicating a certain explanatory power for the total motor score of the FIM. Furthermore, adding the TIS score significantly increased the change in F-value from the Step 1 model, indicating that the TIS score contributed to improving the model (ΔF = 4.55, p = 0.04). The results of both hierarchical multiple regression analyses revealed that the combined evaluation of trunk and lower limb functions better reflected ADL independence than did individual evaluations.

## 4 Discussion

In this study, we analyzed the effects of trunk function and lower limb motor paralysis on ADL independence in a stepwise manner, clarifying the extent of their impact on ADL when assessed individually and in combination. The importance of evaluating both functions in combination was also verified. We found that the accuracy in reflecting ADL independence was improved when trunk function and lower limb motor paralysis were evaluated together rather than individually. This result suggests that it is important to evaluate trunk and lower limb functions in combination for rehabilitation of patients with stroke.

The study revealed an increase in the change in F-value when both functions were added to the regression model versus when either trunk function or lower limb motor paralysis was used individually; thus, the combination of functions improved the explanatory power of the model. This may be because the ADL of patients with stroke involve a variety of movements performed in sitting and standing positions. The FIM includes items such as toileting, transfers, mobility, and stair climbing, all of which require balance and mobility in a standing position (Granger et al., 1993).

While trunk function is crucial for sitting balance, lower limb function—particularly the strength of the muscles around the hip joint—is important for standing balance and walking (Kirker et al., 2000; Cabanas-Valdés et al., 2013). Therefore, practical ADL requires both trunk and lower limb functions. This suggests that combining the evaluation of trunk function and lower limb motor paralysis better explains ADL independence than do individual evaluations.

Furthermore, both the lateral and medial motor systems play essential roles in the acquisition of ADL movements after stroke (Ishiwatari et al., 2021). The lateral motor system mainly controls the distal muscles of the limbs via the corticospinal tract, while the medial motor system controls the trunk and proximal muscles of the limbs via the corticoreticulospinal and corticorubral tracts. These neural pathways are closely situated, rendering them highly susceptible to simultaneous damage during stroke (Jang and Lee, 2019). From the perspective of the neural fibers involved, this suggests the need to comprehensively evaluate both trunk and lower limb functions.

The relationship between trunk and lower limb functions has been reported in previous studies. Verheyden et al. have shown that trunk stability is a prerequisite for the movement of the head and limbs and that these functions are related to ADL (Verheyden et al., 2007). Hsieh et al. have also investigated the impact of walking ability after stroke on ADL, reporting that the recovery of lower limb function is directly linked to improvements in ADL independence (Hsieh et al., 2002). Based on the results of the present study, it was found that in post-stroke rehabilitation, a combined evaluation of both trunk and lower limb function better reflected ADL independence than did evaluating these functions individually. Therefore, to assess the effectiveness of rehabilitation and understand the physical condition, it may be useful to comprehensively evaluate the relationship between trunk function and lower limb motor paralysis.

The novelty of this study lies in the fact that it addresses the insufficient consideration of the interaction between trunk and lower limb functions in previous research. It also complements previous studies that have focused solely on trunk function by means of a detailed analysis with practical ADL evaluations (Hsieh et al., 2002; Sullivan et al., 2002). Herein, hierarchical multiple regression analysis was performed using the FIM—which measures practical ADL independence—as the dependent variable, with trunk function and lower limb motor paralysis as independent variables, providing new insights into the influence of these functions on ADL independence.

The limitations of this study include its relatively small sample size; hence, caution should be exercised when generalizing the results. A larger sample size would enable the construction of more accurate models and improve the reproducibility of the results. Additionally, the cross-sectional design precluded the evaluation of long-term rehabilitation outcomes. Longitudinal follow-up is required to clarify how the effects of rehabilitation change over time. Moreover, the selection of clinical assessment tools used in this study was limited—to more comprehensively evaluate ADL independence, it may be necessary to assess not only motor paralysis but also strength, sensory function, and coordination.

In conclusion, we investigated the impact of evaluating trunk function and lower limb motor paralysis individually and in combination on explaining ADL independence in patients with stroke. The combined evaluation method, which assesses both trunk function and lower limb motor paralysis, more accurately reflected ADL independence, suggesting its usefulness as an assessment method. Future studies should include larger sample sizes, incorporate a longitudinal design, and adopt more multifaceted evaluation methods to examine the effects of trunk and lower limb functions on ADL from multiple perspectives and address the limitations of our study.

## Data Availability

The data that support the findings of this study are available on request from the corresponding author, H.N. The data are not publicly available due to their containing information that could compromise the privacy of research participants.

## 6 Author Contributions

Conceptualization, T.N., and H.N.; Methodology, T.N., and H.N.; Formal Analysis, T.N., and S.S.; Investigation, T.N.; Resources, T.N., and H.N.; Data Curation, T.N.; Writing—Original Draft Preparation, T.N.; Writing—Review and Editing, T.N., S.S., S.F., R.Y., Y.S., T.M., K.N., and H.N.; Visualization, T.N.; Supervision, H.N.; Project Administration, T.N., and H.N.; Funding Acquisition, T.M., K.N., and H.N. All authors have read and agreed to the published version of the manuscript.

## 8 Acknowledgments

We would like to thank the patients who participated in this study.

## 9 Data Availability Statement

The data that support the findings of this study are available on request from the corresponding author, HN, upon reasonable request. The data are not publicly available due to their containing information that could compromise the privacy of research participants.

## Reference

Brunnstrom, S. (1966). Motor testing procedures in hemiplegia: based on sequential recovery stages. Phys. Ther. 46, 357–375.

Cabanas-Valdés, R., Cuchi, G. U., and Bagur-Calafat, C. (2013). Trunk training exercises approaches for improving trunk performance and functional sitting balance in patients with stroke: a systematic review. NeuroRehabilitation 33, 575–592.

Du, M., Mi, D., Liu, M., and Liu, J. (2023). Global trends and regional differences in disease burden of stroke among children: a trend analysis based on the global burden of disease study 2019. BMC Public Health 23, 2120.

Feigin, V. L., Brainin, M., Norrving, B., Martins, S., Sacco, R. L., Hacke, W., et al. (2022). World stroke organization (WSO): Global Stroke Fact Sheet 2022. Int. J. Stroke 17, 18–29.

Fugl-Meyer, A. R., Jääskö, L., Leyman, I., Olsson, S., and Steglind, S. (1975). The post-stroke hemiplegic patient. 1. a method for evaluation of physical performance. Scand. J. Rehabil. Med. 7, 13–31.

Gowland, C., Stratford, P., Ward, M., Moreland, J., Torresin, W., Van Hullenaar, S., et al. (1993). Measuring physical impairment and disability with the Chedoke-McMaster Stroke Assessment. Stroke 24, 58–63.

Granger, C. V., Hamilton, B. B., Linacre, J. M., Heinemann, A. W., and Wright, B. D. (1993). Performance profiles of the functional independence measure. Am. J. Phys. Med. Rehabil. 72, 84–89.

Hsieh, C.-L., Sheu, C.-F., Hsueh, I.-P., and Wang, C.-H. (2002). Trunk control as an early predictor of comprehensive activities of daily living function in stroke patients. Stroke 33, 2626–2630.

Ishiwatari, M., Honaga, K., Tanuma, A., Takakura, T., Hatori, K., Kurosu, A., et al. (2021). Trunk impairment as a predictor of activities of daily living in acute stroke. Front. Neurol. 12, 665592.

Jang, S. H., and Lee, S. J. (2019). Corticoreticular tract in the human brain: A mini review. Front. Neurol. 10, 1188.

Karatas, M., Cetin, N., Bayramoglu, M., and Dilek, A. (2004). Trunk muscle strength in relation to balance and functional disability in unihemispheric stroke patients. Am. J. Phys. Med. Rehabil. 83, 81–87.

Kirker, S. G., Jenner, J. R., Simpson, D. S., and Wing, A. M. (2000). Changing patterns of postural hip muscle activity during recovery from stroke. Clin. Rehabil. 14, 618–626.

Kwakkel, G., Kollen, B. J., and Wagenaar, R. C. (2002). Long term effects of intensity of upper and lower limb training after stroke: a randomised trial. J. Neurol. Neurosurg. Psychiatry 72, 473–479.

Liu, R., and Liu, J. (2022). Prognostic factors of functional outcome in post-acute stroke in the rehabilitation unit. J. Formos. Med. Assoc. 121, 568–569.

Matsuyama, A. (2018). Factors associated with the walking ability of hemiplegic stroke patients. Open J. Nurs. 08, 14–25.

Meijer, R., Ihnenfeldt, D. S., de Groot, I. J. M., van Limbeek, J., Vermeulen, M., and de Haan, R. J. (2003). Prognostic factors for ambulation and activities of daily living in the subacute phase after stroke. A systematic review of the literature. Clin. Rehabil. 17, 119–129.

Patel, A. T., Duncan, P. W., Lai, S. M., and Studenski, S. (2000). The relation between impairments and functional outcomes poststroke. Arch. Phys. Med. Rehabil. 81, 1357–1363.

Pradon, D., Roche, N., Enette, L., and Zory, R. (2013). Relationship between lower limb muscle strength and 6-minute walk test performance in stroke patients. J. Rehabil. Med. 45, 105–108.

Sullivan, K. J., Knowlton, B. J., and Dobkin, B. H. (2002). Step training with body weight support: effect of treadmill speed and practice paradigms on poststroke locomotor recovery. Arch. Phys. Med. Rehabil. 83, 683–691.

Thijs, L., Voets, E., Denissen, S., Mehrholz, J., Elsner, B., Lemmens, R., et al. (2023). Trunk training following stroke. Cochrane Database Syst. Rev. 3, CD013712.

Van Criekinge, T., Saeys, W., Hallemans, A., Vereeck, L., De Hertogh, W., Van de Walle, P., et al. (2017). Effectiveness of additional trunk exercises on gait performance: study protocol for a randomized controlled trial. Trials 18, 249.

Van Criekinge, T., Truijen, S., Schröder, J., Maebe, Z., Blanckaert, K., van der Waal, C., et al. (2019). The effectiveness of trunk training on trunk control, sitting and standing balance and mobility post-stroke: a systematic review and meta-analysis. Clin. Rehabil. 33, 992–1002.

van der Putten, J. J., Hobart, J. C., Freeman, J. A., and Thompson, A. J. (1999). Measuring change in disability after inpatient rehabilitation: comparison of the responsiveness of the Barthel index and the Functional Independence Measure. J. Neurol. Neurosurg. Psychiatry 66, 480–484.

Verheyden, G., Nieuwboer, A., De Wit, L., Feys, H., Schuback, B., Baert, I., et al. (2007). Trunk performance after stroke: an eye catching predictor of functional outcome. J. Neurol. Neurosurg. Psychiatry 78, 694–698.

Verheyden, G., Nieuwboer, A., Mertin, J., Preger, R., Kiekens, C., and De Weerdt, W. (2004). The Trunk Impairment Scale: a new tool to measure motor impairment of the trunk after stroke. Clin. Rehabil. 18, 326–334.

Verheyden, G., Vereeck, L., Truijen, S., Troch, M., Herregodts, I., Lafosse, C., et al. (2006). Trunk performance after stroke and the relationship with balance, gait and functional ability. Clin. Rehabil. 20, 451–458.

Wang, C.-H., Hsueh, I.-P., Sheu, C.-F., and Hsieh, C.-L. (2005). Discriminative, predictive, and evaluative properties of a trunk control measure in patients with stroke. Phys. Ther. 85, 887– 894.

Zhang, X., Ye, W.-Q., Xin, X.-K., Gao, Y.-J., and Yang, F. (2024). Global, regional, and national burden of stroke attributable to diet high in sodium from 1990 to 2019: a systematic analysis from the global burden of disease study 2019. Front. Neurol. 15, 1437633.

